# Gender-affirming care, mental health, and economic stability in the time of COVID-19: a global cross-sectional study of transgender and non-binary people

**DOI:** 10.1101/2020.11.02.20224709

**Authors:** Brooke A. Jarrett, Sarah M. Peitzmeier, Arjee Restar, Tyler Adamson, Sean Howell, Stefan Baral, S. Wilson Beckham

## Abstract

**Background:** Transgender and non-binary people are disproportionately burdened by barriers to quality healthcare, mental health challenges, and economic hardship. This study examined the impact of the novel coronavirus disease (COVID-19) pandemic and subsequent control measures on gender-affirming care, mental health, and economic stability among transgender and non-binary people globally.

**Methods:** We collected global cross-sectional data from 964 transgender and non-binary adult users of the Hornet and Her apps from April to August 2020 to characterize changes in gender-affirming care, mental health, and economic stability as a result of the COVID-19 pandemic. We conducted Poisson regression models to assess if access to gender-affirming care and ability to live according to one’s gender were related to depressive symptoms, anxiety, and changes in suicidal ideation.

**Results:** Individuals resided in 76 countries, including Turkey (27.4%,n=264/964) and Thailand (20.6%,n=205). A majority were non-binary (66.8%,n=644) or transfeminine (29.4%,n=283). Due to the COVID-19 pandemic, 55.0% (n=320/582) reported reduced access to gender-affirming resources, and 38.0% (n=327/860) reported reduced time lived according to their gender. About half screened positive for depression (50.4%,442/877) and anxiety (45.8%,n=392/856). One in six (17.0%,n=112/659) expected losses of health insurance, and 77.0% (n=724/940) expected income reductions. The prevalence of depressive symptoms, anxiety, and increased suicidal ideation were 1.63 (95% CI: 1.36-1.97), 1.61 (95% CI: 1.31-1.97), and 1.74 (95% CI: 1.07-2.82) times higher for individuals whose access to gender-affirming resources was reduced versus not.

**Discussion:** The COVID-19 pandemic has reduced access to gender-affirming resources and the ability of transgender and non-binary people to live according to their gender worldwide. These reductions may drive the increased depressive symptoms, anxiety, and suicidal ideation reported in this sample. To improve transgender and non-binary health globally, increased access to gender-affirming resources should be achieved through policies (e.g., digital prescriptions), flexible interventions (e.g., telehealth), and support for existing transgender health initiatives.

## INTRODUCTION

The global pandemic caused by the SARS-CoV-2 virus has resulted in more than 37 million cases of novel coronavirus disease (COVID-19) and over one million deaths.^1^ In response, countries have implemented a wide range of measures to quell its spread — shelter-in-place orders, closures of business and schools, and the cancellation of surgeries perceived to be elective.^2^ While these interventions are focused on reducing COVID-19 cases and increasing healthcare capacity, they have also negatively affected healthcare access, mental health, and economic stability worldwide.^3–6^

A growing body of literature describes global interruptions to prescriptions for diseases like HIV, increases in depression, and significant losses of job, insurance, and food security as a result of the COVID-19 pandemic.^7–11^ However, recent work has demonstrated that these effects disproportionately burden groups that have been historically marginalized. Specifically, pandemic control measures have exacerbated existing health disparities and social inequities along lines of poverty and occupation, race and ethnicity, and sexual orientation.^12–16^ The COVID-19 pandemic may also uniquely affect transgender and non-binary people.

Prior to the COVID-19 pandemic, transgender and non-binary individuals experienced barriers to care, greater mental health challenges, and economic vulnerabilities caused by stigma, discrimination, and minority stress.^17–19^ Transgender and non-binary populations face a scarcity of clinicians trained in gender-affirming practices and widespread transphobia among healthcare staff, both of which make healthcare less accessible.^20–22^ Yet access to gender-affirming healthcare (e.g., chest/breast surgery), services (e.g., hair removal), and goods (e.g., binders and packers) can substantially improve the quality of life and mental health of transgender and non-binary populations, who frequently have elevated levels of depression, anxiety, and suicidal ideation.^23–26^ A disproportionate number of transgender and non-binary individuals also experience structural vulnerabilities, such as economic, food, and housing insecurity, that can reinforce or worsen barriers to gender-affirming resources and mental health counseling.^27,28^

The COVID-19 pandemic is exacerbating these risks among transgender and non-binary individuals. For example, there have been documented cancellations and delays in gender-affirming surgeries;^29^ such delays and cancellations have previously been connected to negative mental health consequences.^23,30^ Furthermore, many transgender and non-binary individuals who were living according to their gender prior to the COVID-19 pandemic have had to return to living according to their sex assigned at birth upon moving in with relatives.^31^ Researchers have also reported on gendered policies from Panama, Peru, and Columbia that attempted to reduce the density of crowds in public places by requiring women and men to access essential services on alternating days — a policy that, like other gender-based laws, would likely result in violence against transgender communities.^32^ In these situations, transgender and non-binary individuals may be limited in their ability to live safely and comfortably as themselves. The impact of the COVID-19 pandemic is likely to be especially adverse for those who are already economically marginalized, occupying other marginalized identities (e.g., people who are racial or ethnic minorities, living with HIV, or living with disabilities), or both.

Of the studies to date about the impact of COVID-19 on transgender and non-binary individuals, a majority have been conducted in a single country like the United States and focused on measuring a narrow spectrum of indicators. The objective of this study was to describe the global impact of the COVID-19 pandemic and subsequent control measures on gender-affirming care, mental health, and economic stability among transgender and non-binary individuals. We also examined the association between reduced access to gender-affirming care and ability to live according to one’s gender with multiple mental health indicators.

## METHODS

### Study Design and Participants

For this cross-sectional study, we used data from the global COVID-19 Disparities Survey distributed between April 16 and August 3, 2020 via Hornet and Her — social networking apps marketed to sexual minorities, both cisgender and transgender. We sent survey invitations to the app-specific inboxes of all users who had been active on their app for at least one year. To take the survey, individuals had to report being 18 years or older, view a consent form, and indicate their informed consent by clicking a button to begin.

A total of 24,618 individuals began the survey. For these analyses, we included 1,285 transgender and non-binary adults, which we defined as people 18 years or older who self-reported being transgender, non-binary, or a gender different than their sex assigned at birth. We excluded all women assigned female or intersex at birth (n=161), all men assigned male or intersex at birth (n=12,740), individuals who did not report a gender (n=9,751), individuals who only reported not knowing, not wishing to disclose, or being unable to disclose their gender (n=654), and non-transgender identifying men and women who did not report an assigned sex at birth (n=27).

To ensure the quality of our study population, we screened for duplicate survey responses based on IP address, and again by searching for identical responses to twenty randomly selected variables but found none. We also excluded individuals who completed 89% or less of the survey (n=271), who finished in less than the minimum piloted time of seven minutes (n=47), or provided conflicting responses for multiple questions (n=3) for a final sample size of 964. The Johns Hopkins School of Public Health Institutional Review Board provided a Category 4 exemption to the larger survey prior to distribution.

### Demographic Measures

Individuals self-reported gender by choosing any number of the following options: woman, man, transgender man, transgender woman, or non-binary (including gender-diverse, genderqueer, gender nonconforming, gender expansive, and agender). They also self-reported their country of residence, age, socioeconomic status, years of education, ethnic minority and immigration status, access to masks during the COVID-19 pandemic, and whether the government in their area had ever imposed confinement orders (e.g., mobility restrictions to stay-at-home). We categorized countries according to regions defined by the World Health Organization.

We used eight mutually exclusive categories to describe reported genders. To increase the power for our analyses, we then collapsed individuals into three groups, building on recommendations from Reisner et al: (1) transmasculine, i.e., people who were assigned female at birth (AFAB) or intersex who self-reported being transgender or being a man; (2) transfeminine, i.e., people who were assigned male at birth (AMAB) or intersex who self-reported being transgender or being a woman; and (3) non-binary, i.e., individuals who reported being either solely non-binary, both a man and a woman, or a transgender man and transgender woman.

We chose to operationalize three gender categories for multiple reasons. First, while some non-binary individuals explicitly reported also being transgender, the majority did not. We wanted to honor this distinction while also allowing for individuals who reported being both men and women to transcend the transfeminine versus transmasculine binary. Second, our survey presented a limited number of gender options to a global audience in which being a third gender (e.g., two-spirit, bissu, fa’afafine, qariwarmi) is distinct from many Western concepts of being transgender ^33^. Lastly, in line with recommendations from Restar et al., we saw statistically significant differences when comparing non-binary individuals with transmasculine and transfeminine individuals ^34^. Therefore, presenting results stratified by gender (i.e., a “gender-specific” approach) was more appropriate than conducting analyses on all individuals together and presenting them as a single population (i.e., a “gender-inclusive” approach).

### COVID-19 Impact Measures

Individuals answered questions about the impact of the COVID-19 pandemic and the resulting response on their access to gender-affirming resources, their mental health, and their economic stability. For indicators related to gender-affirming resources, we asked those who self-identified as transgender or non-binary (n=865) whether the COVID-19 pandemic limited their access to the following: hormone therapy and/or medications; surgical aftercare materials (e.g., vaginal dilators); cosmetic supplies and services (e.g. makeup, wigs, and hair removal); mental health counseling and therapy services; and body modifiers (e.g., binders and packing materials); to which they could respond, “Yes,” “No,” or “Not Applicable.” We characterized the severity of interruptions to gender-affirming care by whether individuals reported that more than one resource had been impacted. We also asked whether the COVID-19 pandemic had changed the amount of time that the individual could live according to their gender (“Compared to before the COVID-19 crisis, how often are you able to live according to your gender identity?”). We categorized the five-point Likert scale into three categories: More than before (i.e., more or a lot more as compared to before COVID-19), about the same as before COVID-19, and less than before (i.e., less than or not at all as compared to before COVID-19).

For mental health indicators, we used the 4-item patient health questionnaire (PHQ-4) to screen for common symptoms of depression and anxiety, which we dichotomized with a score of three or more being considered positive ^35^. We assessed the impact of COVID-19 on loneliness (“Have you been feeling lonely since the COVID crisis began?”) using a four-point Likert Scale, which we dichotomized into a positive sentiment (“not lonely” or “not much lonely”) and negative sentiment (“very much lonely” or “a little lonely”). We also asked how often they had thought about taking their own life presently and in the six months prior to the COVID-19 pandemic with the following answer options: “never,” “seldom,” “quite often,” “very often,” and “all the time.” We created four categories to describe changes from pre- to mid-pandemic: was and remains rare (i.e., “never” or “seldom”), was and remains frequent (i.e., “quite often,” “very often,” or “all the time”), decreased, or increased. To characterize resiliency, we asked about the following in the face of the COVID-19 pandemic: if they had “sources of hope, strength, comfort, and peace”; if they were “intent on finding emotional support and therapy for themselves”; and if they believed that they were able to “live a happy, full life despite the crisis.”

### Statistical Analyses

We conducted descriptive analyses of demographic measures and presented descriptions of the COVID-19 impact measures stratified by gender, using chi-squared and Fisher’s exact tests as appropriate. We also stratified results by country in the supplementary materials. We used Poisson regression models with complete case analyses to assess for differences in the prevalence of screening positive for depression and anxiety among individuals with reduced (versus continued) access to gender-affirming resources and individuals who reported being able to live according to their gender less (versus more) since the COVID-19 pandemic started. We reported these comparisons as prevalence ratios. We used the same approach to assess the impact of reductions in access to care and changes in ability to live according to one’s gender on changes in suicidal ideation, stratified on baseline levels of suicidal ideation. We conducted all analyses in Stata version 14.^36^

## RESULTS

These analyses primarily consisted of non-binary (66.8%, n=644/964) and transfeminine (29.4%, n=283) individuals (**Table 1**). About 47% (n=451) were from the European region and 25.1% (n=242) were from the South-East Asia region. There were 76 countries represented in the sample; a majority of individuals were residents of Turkey (27.4%, n=264), Thailand (20.6%, n=199), and Russia (11.5%, n=117). No other single country accounted for more than 5% of the sample. Individuals were young and highly educated, with 50.5% (n=487) being between 18 and 29 years old and 42.6% (n=410) having at least a university degree. Few (12.7%, n=122) had ever lacked access to a mask during the COVID-19 pandemic and 75.6% (n=729) lived in a country that had issued COVID-related confinement or “stay-at-home” orders.

**Table 1:** Demographics for transgender and non-binary individuals from the COVID-19 Disparities Survey distributed globally by the Hornet and Her apps between April 16 and August 3, 2020 (N=964)

|  | Overall (%)<sup>a</sup> |
| --- | --- |
| **Self-Reported Transgender Identity** |  |
| Transgender man | 95 (9.9%) |
| Transgender woman | 201 (20.9%) |
| Non-binary (NB) only | 594 (61.6%) |
| NB transgender woman | 17 (1.8%) |
| NB transgender man | 7 (0.7%) |
| NB, transgender woman, and transgender man | 2 (0.2%) |
| Transgender man and transgender woman | 7 (0.7%) |
| Man and woman | 41 (4.3%) |
| **Researcher-Generated Gender Categories** |  |
| Transmasculine (assigned female sex or intersex at birth) | 37 (3.8%) |
| Transfeminine (assigned male sex or intersex at birth) | 283 (29.4%) |
| Non-binary<sup>b</sup> | 644 (66.8%) |
| **World Health Organization Region** |  |
| Europe | 451 (46.8%) |
| South-East Asia | 242 (25.1%) |
| Americas | 86 (8.9%) |
| Eastern Mediterranean | 85 (8.8%) |
| Western Pacific | 40 (4.2%) |
| Africa | 35 (3.6%) |
| **Age** |  |
| 18-29 years | 487 (50.5%) |
| 30 - 39 years | 287 (29.8%) |
| 40 - 49 years | 132 (13.7%) |
| 50+ years | 58 (6.0%) |
| **Socioeconomic Status** |  |
| Lower | 151 (15.7%) |
| Lower Middle | 472 (49.0%) |
| Upper middle | 290 (30.1%) |
| Upper | 46 (4.8%) |
| **Years of Education** |  |
| Less than 6 years | 54 (5.6%) |
| 6-12 years | 176 (18.3%) |
| Some university, no degree | 193 (20.0%) |
| Trade school | 120 (12.5%) |
| University degree or more | 410 (42.6%) |
| **Ethnic Minority** |  |
| Yes | 250 (26.0%) |
| No | 493 (51.2%) |
| Don't know / can't answer | 211 (21.9%) |
| **Immigrant** |  |
| Yes | 143 (14.8%) |
| No | 719 (74.6%) |
| Not sure | 85 (8.8%) |
| **COVID-19 Pandemic Environment** |  |
| Ever lacked access to a mask | 122 (12.7%) |
| In a location that ever issued "stay-at-home" confinement orders | 729 (75.6%) |

<sup>a</sup> Denominators include those for whom data were missing: 25 did not report a country, 5 did not report a socioeconomic status, 11 did not report an education level, 10 did not report if they were an ethnic minority, 17 did not report if they were an immigrant, 6 did not report their access to masks, and 7 did not report whether their country of residence had implemented "stay-at-home" confinement orders <sup>b</sup> Of those whose gender was non-binary, 28 reported being assigned female sex at birth, 578 reported being assigned male sex at birth, 37 reported being assigned intersex at birth, and one did not report their sex assigned at birth.

More than half (55%, n=320/582) of the sample reported that the COVID-19 pandemic had limited their access to one or more gender-affirming resource (**Table 2**). Mental health counseling and therapy was the most commonly cited resource to be affected (42.9%, n=192/448), with a somewhat greater proportion of transmasculine individuals reporting reduced access to counseling (61.9%, n=13/21) than non-binary (43.0%, n=122/284) and transfeminine (39.9%, n=57/143) individuals (p-value=0.16). Transmasculine and transfeminine individuals were more likely than non-binary individuals to report that the COVID-19 pandemic limited their access to gender-affirming hormones and medications (55.0% [n=11/20] and 42.1% [n=61/145] vs. 30.1% [n=71/236], p-value=0.01) as well as surgical aftercare materials (42.9% [n=6/14] and 40.2% [n=51/127] vs. 28.8% [n=62/215], p-value=0.08). All geographic regions reported reductions in access to gender-affirming resources; at least half of the individuals in each region, beside the Western Pacific, reported reduced access to one or more resource (**S1 Table**). More than a third (38.0%, n=327/860) of individuals reported that the COVID-19 pandemic had reduced or completely eliminated their ability to live according to their gender, with more transfeminine individuals (43.1%, n=100/232) being unable to living according to their gender as compared to transmasculine (28.6%, n=8/28) and non-binary (36.5%, n=219/600) individuals (p-value <0.001).

**Table 2:** Access to and actualization of gender-affirming resources among self-identified transgender and non-binary individuals from the COVID-19 Disparities Survey between April 16, 2020 and August 3, 2020 (N=964)

|  | Overall (%) | Transmasculine | Transfeminine | Non-Binary | p-value<sup>b</sup> |
| --- | --- | --- | --- | --- | --- |
| Experienced reduced access to one or more gender-affirming resource below<sup>a</sup> | 320 / 582<br>(55.0%) | 17 / 26<br>(65.4%) | 110 / 189<br>(58.2%) | 193 / 367<br>(52.6%) | 0.25 |
| Hormone therapy and/or gender-affirming medication | 143 / 401<br>(35.7%) | 11 / 20<br>(55.0%) | 61 / 145<br>(42.1%) | 71 / 236<br>(30.1%) | 0.01 |
| Surgical aftercare | 119 / 356<br>(33.4%) | 6 / 14<br>(42.9%) | 51 / 127<br>(40.2%) | 62 / 215<br>(28.8%) | 0.08 |
| Cosmetic supplies and services, e.g., makeup, wigs, and hair removal | 189 / 500<br>(37.8%) | 9 / 21<br>(42.9%) | 65 / 162<br>(40.1%) | 115 / 317<br>(36.3%) | 0.63 |
| Mental health counseling and therapy* | 192 / 448<br>(42.9%) | 13 / 21<br>(61.9%) | 57 / 143<br>(39.9%) | 122 / 284<br>(43.0%) | 0.16 |
| Body modifiers, e.g., binders and packing material | 160 / 443<br>(36.1%) | 8 / 18<br>(44.4%) | 57 / 148<br>(38.5%) | 95 / 277<br>(34.3%) | 0.52 |
| Compared to before the COVID-19 pandemic, how often able to live according to their gender |  |  |  |  |  |
| More or a lot more | 67 / 860<br>(7.8%) | 7 / 28<br>(25.0%) | 22 / 232<br>(9.5%) | 38 / 600<br>(7.1%) | < 0.001 |
| About the same | 466<br>(54.2%) | 13<br>(46.4%) | 110<br>(47.4%) | 343<br>(57.2%) |  |
| Less or not at all | 327<br>(38.0%) | 8<br>(28.6%) | 100<br>(43.1%) | 219<br>(36.5%) |  |

<sup>a</sup> Denominators excluded participants who were not presented with these questions, did not respond, or said that the resource was not applicable to them
<sup>b</sup> p-values were calculated using chi-squared and Fischer's exact tests as appropriate

About half of the sample screened positive for depression (50.4%, n=442/877) and anxiety (45.8%, n=392/856), with a larger proportion of transfeminine individuals reporting these outcomes than transmasculine and non-binary individuals (**Table 3**). Overall, 10.0% (n=93/928) reported that suicidal ideation had increased during the COVID-19 pandemic, and 12.5% (n=116) reported that it had decreased. Transfeminine individuals were more likely to report increases in suicidal ideation (11.6%, n=31/263) while being less likely to agree with statements of resiliency, such as having sources of hope, strength, comfort, and peace (47.9%, n=100/209) when contrasted with transmasculine (8.3%, n=3/36; 71.0%, n=22/31) and non-binary (9.5%, n=59/624; 69.5%, n=367/528) individuals (p-value=0.98; p-value < 0.001). Sixteen percent (n=136/851) of individuals reported that they were not intent on finding emotional support and therapy for themselves during the COVID-19 pandemic. Seventy-seven percent (n=724/940) of the sample expected a reduction in their income, 17% (n=112/659) expected to lose health insurance, and 53.4% (n=428/801) reported not having received financial aid, despite need (**Table 4**). Though 40% (n=361/900) of individuals overall reported cutting or reducing their meals, fewer non-binary individuals had done so (34.9% [n=212/607] versus 51.4% [n=18/35] of transmasculine individuals and 50.8% [n=131/258] of transfeminine individuals).

**Table 3:** Mental health and resiliency indicators among transgender and non-binary individuals from the COVID-19 Disparities Survey between April 16, 2020 and August 3, 2020 (N=964) ^a^

|  | Overall (%) | Transmasculine | Transfeminine | Non-Binary | p-value<sup>b</sup> |
| --- | --- | --- | --- | --- | --- |
| Screened positive (PHQ-4 ≥3) |  |  |  |  |  |
| Depression | 442 / 877<br>(50.4%) | 17 / 35<br>(48.6%) | 144 / 245<br>(58.8%) | 281 / 597<br>(47.1%) | 0.01 |
| Anxiety | 392 / 856<br>(45.8%) | 17 / 33<br>(51.5%) | 125 / 237<br>(52.7%) | 250 / 586<br>(42.7%) | 0.03 |
| Felt lonely since COVID-19 began |  |  |  |  |  |
| Yes | 685 / 957<br>(71.6%) | 28 / 37<br>(75.7%) | 216 / 281<br>(76.9%) | 441 / 639<br>(69.0%) | 0.04 |
| Frequency of suicidal ideation since COVID-19 vs. 6 months prior |  |  |  |  |  |
| Was and remains rare | 648 / 928<br>(69.8%) | 26 / 36<br>(72.2%) | 183 / 268<br>(68.3%) | 439 / 624<br>(70.4%) | 0.98 |
| Decreased from frequent to rare | 116<br>(12.5%) | 4<br>(11.1%) | 33<br>(12.3%) | 79<br>(12.7%) |  |
| Increased from rare to frequent | 93<br>(10.0%) | 3<br>(8.3%) | 31<br>(11.6%) | 59<br>(9.5%) |  |
| Was and remains frequent | 71<br>(7.6%) | 3<br>(8.3%) | 21<br>(7.8%) | 47<br>(7.5%) |  |
| Reported having sources of hope, strength, comfort, and peace |  |  |  |  |  |
| Yes | 489 / 768<br>(63.7%) | 22 / 31<br>(71.0%) | 100 / 209<br>(47.9%) | 367 / 528<br>(69.5%) | < 0.001 |
| Reported being intent on finding emotional support and therapy<sup>c</sup> |  |  |  |  |  |
| Agree | 467 / 851<br>(54.9%) | 20 / 35<br>(57.1%) | 129 / 237<br>(54.4%) | 318 / 579<br>(54.9%) | 0.85 |
| Disagree | 136<br>(16.0%) | 5<br>(14.3%) | 43<br>(18.1%) | 88<br>(15.2%) |  |
| Reported believing they could live a happy, full life despite the pandemic<sup>c</sup> |  |  |  |  |  |
| Agree | 516 / 847<br>(60.9%) | 26 / 34<br>(76.5%) | 137 / 231<br>(59.3%) | 353 / 582<br>(60.6%) | 0.22 |
| Disagree | 120<br>(14.2%) | 2<br>(5.9%) | 40<br>(17.3%) | 78<br>(13.4%) |  |

<sup>a</sup> Denominators excluded individuals who did not respond or reported not knowing their answer unless otherwise noted <sup>b</sup> p-values were calculated using chi-squared and Fischer's exact tests as appropriate <sup>c</sup> Denominator includes those who stated "neither agree nor disagree"

**Table 4:** Economic indicators among transgender and non-binary individuals from the COVID-19 Disparities Survey between April 16, 2020 and August 3, 2020 (N=964) ^a^

|  | Overall (%) | Transmasculine | Transfeminine | Non-Binary | p-value<sup>b</sup> |
| --- | --- | --- | --- | --- | --- |
| Lost job due to COVID-19 | 151 / 953<br>(15.8%) | 5 / 37<br>(13.5%) | 53 / 278<br>(19.1%) | 93 / 638<br>(14.6%) | 0.21 |
| Expected reduction in income |  |  |  |  |  |
| 0% | 216 / 940<br>(23.0%) | 13 / 37<br>(35.1%) | 57 / 270<br>(21.1%) | 10 / 633<br>(23.1%) | 0.33 |
| 1-39% | 265<br>(28.2%) | 11<br>(29.7%) | 70<br>(25.9%) | 184<br>(29.1%) |  |
| 40-99% | 337<br>(35.8%) | 9<br>(24.3%) | 101<br>(37.4%) | 227<br>(35.9%) |  |
| 100% | 122<br>(13.0%) | 4<br>(10.8%) | 42<br>(15.6%) | 76<br>(12.0%) |  |
| Expected to lose health insurance<sup>c</sup> |  |  |  |  |  |
| Yes | 112 / 659<br>(17.0%) | 7 / 23<br>(30.4%) | 40 / 179<br>(22.4%) | 65 / 457<br>(14.2%) | 0.05 |
| No | 408<br>(61.9%) | 11<br>(47.8%) | 104<br>(58.1%) | 293<br>(64.1%) |  |
| Financial Aid |  |  |  |  |  |
| Received and not needed | 22 / 801<br>(2.8%) | 3 / 30<br>(10.0%) | 8 / 229<br>(3.5%) | 11 / 542<br>(2.0%) | 0.08 |
| Not received but not needed | 159<br>(19.8%) | 9<br>(30.0%) | 37<br>(16.2%) | 113<br>(20.8%) |  |
| Received and needed | 192<br>(24.0%) | 5<br>(16.7%) | 56<br>(24.4%) | 131<br>(24.2%) |  |
| Not received and needed | 428<br>(53.4%) | 13<br>(43.3%) | 128<br>(55.9%) | 287<br>(53.0%) |  |
| Had cut or reduced meals | 361 / 900<br>(40.1%) | 18 / 35<br>(51.4%) | 131 / 258<br>(50.8%) | 212 / 607<br>(34.9%) | < 0.001 |
| Among those employed, ability to miss work |  |  |  |  |  |
| Was telecommuting or on paid leave | 163 / 473<br>(34.5%) | 8 / 23<br>(34.8%) | 32 / 128<br>(25.0%) | 123 / 322<br>(38.2%) | 0.06 |
| Cannot afford to miss work but was following confinement orders | 146<br>(30.9%) | 7<br>(30.4%) | 40<br>(31.2%) | 99<br>(30.8%) |  |
| Cannot afford to stay home and must work to survive | 164<br>(34.7%) | 8<br>(34.8%) | 56<br>(43.8%) | 100<br>(31.1%) |  |

<sup>a</sup> Denominators excluded individuals who did not respond or reported not knowing their answer unless otherwise noted <sup>b</sup> p-values were calculated using chi-squared and Fischer's exact tests as appropriate <sup>c</sup> Denominator includes those who reported they "might or might not" lose their health insurance

A positive screen for depression was 1.63 (95% confidence interval [CI]: 1.36-1.97) times more common among those who had lost access to one or more gender-affirming resource during the COVID-19 pandemic as compared to those without reductions in access (**Table 5**). Similarly, a positive screen for anxiety was 1.61 (95% CI: 1.31-1.97) times more common among those who had lost access to one or more gender-affirming resource compared to those without reductions in access. Trends were similar for suicidal ideation. For example, among those with rare or no suicidal ideation at the beginning of the COVID-19 pandemic, individuals who had reduced access to gender-affirming resources were 1.74 (95% CI: 1.07, 2.82) times more likely to report increased suicidal ideation. Screening positive for depression and anxiety were also 1.21 (95% CI: 0.92, 1.58) and 1.48 (95% CI: 1.04, 2.10) times more prevalent among those reporting that the COVID-19 pandemic had decreased the amount of time they could live according to their gender, versus those who had increased that time. However, among individuals who had no or rare suicidal ideation prior to the COVID-19 pandemic, a smaller proportion of those living less according to their gender during the COVID-19 pandemic had increased suicidal ideation compared to those who lived more as their gender (prevalence ratio = 0.57 [95% CI: 0.33- 0.98]).

**Table 5:** Bivariate prevalence ratios of screening positive for depression, screening positive for anxiety, and changes in suicidal ideation among transgender and non-binary individuals from the COVID-19 Disparities Survey between April 16, 2020 and August 3, 2020

|  | Screening positive<br>for depression<br>PrR (95% CI)<sup>a</sup> | Screening positive<br>for anxiety<br>PrR (95% CI) | Increased frequency<br>of suicidal thoughts<br>(vs. remained low)<sup>b</sup> | Retained frequent<br>suicidal thoughts<br>(vs. reduced frequency)<sup>c</sup> |
| --- | --- | --- | --- | --- |
| Reported reduction in access<br>to more than one (vs. 0)<br>gender affirming resource | 1.63 (1.36, 1.97)<br>n = 532 | 1.61 (1.31, 1.97)<br>n = 523 | 1.74 (1.07, 2.82)<br>n = 449 | 1.37 (0.87, 2.15)<br>n = 112 |
| Reported decreased time<br>(vs. increased) lived<br>according to one's gender | 1.21 (0.92, 1.58)<br>n = 346 | 1.48 (1.04, 2.10)<br>n = 331 | 0.57 (0.33, 0.98)<br>n = 310 | 1.12 (0.50, 2.54)<br>n = 68 |

<sup>a</sup> Prevalence ratio (PrR) and 95% confidence interval (95% CI) <sup>b</sup> Among individuals who reported never or seldom having suicidal ideation in the six months prior to the pandemic beginning <sup>c</sup> Among individuals who reported having suicidal ideation quite often, very often, and all the time in the sex months prior to the pandemic beginning

## DISCUSSION

This survey provides early insights into the impacts of the COVID-19 pandemic on access to gender-affirming resources, mental health, and economic stability among transgender and non-binary communities globally. Roughly half of individuals reported that the COVID-19 pandemic had restricted their access to gender-affirming resources, and nearly two in five reported the COVID-19 pandemic had negatively impacted their ability to live according to their gender. Screening positive for depression and/or anxiety and increases in suicidal ideation were common, but more so for those who experienced reduced access to gender-affirming resources. Of these resources, counseling and therapy were the most affected by COVID-19, but most people who responded to the survey also reported resiliencies, such as having sources of hope and being intent on finding emotional support. This intent to seek support, however, may be dampened by the fact that many individuals expected reductions in income and loss of health insurance. This is the first empirical study to examine the effect of COVID-19 and its impact on gender-affirming resources, mental health, and economic stability among a global, multi-region sample of transgender and non-binary individuals.

Half of the individuals in our survey reported reduced access to one or more gender-affirming resource. This was higher in our sample than in an online survey of 1,240 transgender and non-binary individuals in Germany, Switzerland, and Austria.^29^ A third or more of individuals from the European region in our study reported reductions in access to hormone therapy and surgical aftercare materials, which was higher than the 18% and 3% of individuals from the European-specific survey respectively. That survey also found about a quarter of individuals had delayed or cancelled aftercare for a recent surgery. Differences may be due to the populations sampled, as our survey primarily drew from countries in Eastern Europe where there is substantial stigma against and policing of transgender individuals. Together, though, these results signify that the COVID-19 pandemic is causing decreases in access to gender-affirming resources even in high-income settings.

Our study demonstrated that reduced access to gender-affirming resources due to the COVID- 19 pandemic were associated with poorer mental health. Screening positive for depression, screening positive for anxiety, and increased suicidal ideation were more common for those whose access to one or more gender affirming resource had been reduced due to the COVID- 19 pandemic. These results mirror anecdotal data from clinics serving transgender patients in China.^37^ Gender-affirming resources are crucial to transgender and non-binary individuals, as these resources often activate and enhance the interactive process of receiving recognition for their gender, sense of self, and sense of humanity.^38^ Given the abundant literature that gender affirmation results in better mental health and quality of life, these data underscore the importance of ensuring that transgender and non-binary individuals have access to these essential services and products.^23,24,30,39–41^ Access to these resources should also be fortified to avoid negative physical health outcomes. For example, about a third of this sample reported reductions in access to hormones and 17% reported that they expected to lose their health insurance, which may support hormone therapy procurement. These disruptions may force some transgender and non-binary individuals to discontinue hormone therapy or mete out their limited doses to last longer. Sustained interruptions or sub-optimal dosing of hormone therapy have been connected with symptoms of hypogonadism, such as osteoporosis and cardiovascular disease.^42^ Similarly, aftercare for gender-affirming surgeries is critical for avoiding negative physical outcomes like urinary tract hesitancy and needing re-operation, yet a third of individuals in this study reported that they had reduced access to the surgical aftercare that they needed.^43^

A third of individuals reported decreased time lived according to their gender. In comparison, a poll conducted by the Trevor Project in the United States of 600 lesbian, gay, bisexual, transgender, and questioning (LGBTQ) youth (13-19 years) reported that 56% of the transgender and non-binary individuals had reduced their ability to express their LGBTQ identity due to the COVID-19 pandemic.^44^ Youth often attributed needing to move back in with unsupportive caretakers as a reason for not being able to live according to their gender,^31,44^ which may explain the elevated proportion compared to adults in our study. Living less according to one’s gender during the COVID-19 pandemic was associated with screening positive for depression and anxiety. However, a counterintuitive result was found among individuals in our study who had rare suicidal ideation prior to the COVID-19 pandemic and lived more according to their gender during the COVID-19 pandemic. This subset of the sample was more likely to have increased suicidal ideation as compared to individuals who were living according to their gender less. These results, however, may reflect a limitation of our measurement tool. For example, the former group likely had a low baseline ability to live according to their gender pre-pandemic and hence could only increase the amount of time lived according to their gender. The worse mental health in this group may be linked to low pre-pandemic levels of living according to their gender, rather than the recent increase in their ability to do so. Because we did not measure pre-pandemic and current ability to live according to one’s gender separately, we could not control for baseline levels in the model. It is also possible that people who recently began living in their gender more may be subjected to elevated levels of anticipated and experienced stigma that could be driving increased suicidality; prior research has shown discrimination due to gender expression, rather than gender identity itself, to be associated with mental distress.^45–47^

Positive screens for depression and anxiety were correlated with decreases in access to gender-affirming care and decreased time lived according to one’s gender, and were present in nearly half the sample. These data align with results from transgender and non-binary youth from the Trevor Project poll and another sample of 201 young adults (19-25 years) attending college, both from the U.S.^44,48^ Findings in a small study of 15 Latinx trans women in the United States suggest that these poor mental health indicators represent declines as a result of the COVID-19 pandemic rather than just a high baseline prevalence.^49^ These findings are particularly concerning when contrasted against the large proportion of individuals in this and other studies who reported that COVID-19 had decreased their access to mental health therapy and counseling.^44,48^ To mitigate the immediate trauma of the COVID-19 pandemic and potential long-lasting effects, innovative mental health interventions — from remote video and phone therapy to self-help apps — have been emerging, but additional investments are needed, especially to reach those with limited to no access to Internet.^50,51^ Furthermore, our findings support the notion that transmasculine, transfeminine, and non-binary populations are having distinct experiences during the COVID-19 pandemic and should receive gender-specific support.^37^ For example, transmasculine individuals were more likely to report having reduced access to gender-affirming resources but generally reported better mental health outcomes, as well as having sources of strength and comfort. Combined with the fact that transgender and non-binary youth have been more likely to reach out to friends and family than cisgender lesbian, gay, and pansexual youth according to the Trevor Project data, it may be possible to enhance these resiliencies by strengthening and expanding trauma-informed, online peer-to-peer support efforts such as Q Chat Space ^31,44,52^. However, different approaches to transfeminine, transmasculine, and non-binary individuals may be needed.

Lastly, we found that transgender and non-binary communities worldwide are experiencing strains across basic needs such as finances, food, and health insurance, all as a result of COVID-19 pandemic. These strains, in addition to pre-existing economic vulnerabilities, will contribute to even greater barriers to gender-affirming care, mental health counseling, services, and products.^27^ For example, approximately 10% of transgender and non-binary individuals in the U.S. lacked health insurance before the COVID-19 pandemic.^27^ Though some may have health insurance through their employers, transgender and non-binary people are also more likely to be employed in the industries most impacted by business shutdowns.^12^ We found that more than three quarters of the sample expected a reduction in their income, one in six expected to lose their health insurance, and more than half reported needing and not receiving financial aid. These results display the pronounced structural vulnerabilities that shape experiences of transgender and non-binary communities in the current context of the COVID-19 pandemic, and likely contribute further as stressors to mental health. These stressors may climb as transgender and non-binary communities continue to experience worsening economic instability due to the COVID-19 pandemic and associated control measures. Further research is necessary to examine the syndemic impact of COVID-19-related stressors on transgender and non-binary communities, particularly those who are experiencing multiple, intersectional stressors at the individual, interpersonal, and structural levels.

This study had some limitations. While the survey reached individuals from six continents and was available in 13 languages, these data are not a representative sample of transgender and non-binary individuals worldwide. As a survey distributed through a mobile app, participation was limited to individuals with Internet and a smartphone. Individuals in the survey were also generally highly educated. Our survey likely missed highly disadvantaged individuals, and consequently, likely underestimates the true magnitude of impact of COVID-19 on this community. Our study also had a methodological limitation in its categorization of genders — while we aimed to offer an inclusive set of options, we did not capture the full spectrum nor fluidity of genders across the cultural diversity of the countries represented. Yet, our sample does present results from a large number of non-binary individuals, which is a population that has generally been understudied but increasingly recognized as distinct from binary transgender individuals.^53^ Furthermore, the survey only presented questions about access to gender-affirming resources to individuals who self-identified as transgender (e.g., versus those who selected being a man and were AFAB but did self-report as transgender). The next wave of data collection will present the module to all persons whose sex assigned at birth does not match their current gender.

The findings of the current study provide insights for future directions. Namely, future research should expand on this work by identifying protective factors that can be potentially leveraged to buffer the impact of COVID-19 pandemic on gender-affirming resources, mental health, and economic stability. There is also a need to contextualize and understand how transgender and non-binary communities are currently responding to the economic instabilities due to the epidemic, particularly in regions where mandatory stay-at-home orders remain. These restrictions may have led or will lead some transgender and non-binary individuals to turn to more dangerous work for income, unregulated and unmonitored markets for gender-affirming services or goods, or the use of alcohol and other substances to cope. Lastly, given the mobile and online nature of recruitment for this study, researchers should look for feasibility and opportunity with Hornet and other mobile apps as a platform for outreach, programming, and interventions for transgender and non-binary communities in regions where the apps are utilized.

## CONCLUSION

Along with these research implications, our findings suggest the need for multiple programmatic interventions specific to transgender and non-binary populations. Maintaining and increasing secure access to lifesaving gender-affirming resources, mental health services, and economic stability will require backing from both typical and atypical sources — from nonprofit organizations to for-profit companies to academic researchers — both during and after the COVID-19 pandemic. This could include, for example, instrumental support according to community needs (e.g., coordinating food bank deliveries or monetary support for bills) or resource mapping to help transgender and non-binary individuals identify where they can seek pandemic-related relief and aid without stigma or discrimination ^52^. Health insurers and healthcare facilities could also transparently communicate changes in policies and logistics for gender-affirming services to alleviate anxieties around loss of access due to pandemic control measures. To prevent detrimental mental health consequences due to inaccessibility of gender-affirming resources and economic hardships, rapid policies (e.g., digital prescription refills) and flexible interventions (e.g., telehealth) are needed to maintain continuity of gender-affirming hormones as well as therapy and counseling. To achieve these interventions, innovative partnerships will be needed to reach the most marginalized— both by supporting trans-led, community-based organizations to maintain and expand their transgender health services as well as by increasing the capabilities of nation- and/or state-sponsored programs and private sector companies to better serve transgender and non-binary communities.

## Data Availability

Data cannot be shared publicly per an IRB agreement on data sharing to minimize the
risk to the gender minorities participating in this survey and to maximize privacy and
confidentiality as much as possible.

## Acknowledgements

The authors thank the individuals who volunteered their valuable time to participate in our survey. The authors also thank the people who translated the survey from English into 13 different languages: Ana Cara-Linda, Anna Yakusik, Carol Strong, Damiano Cerasuolo, Edward Sutanto, Ezgi Ayeser, Henrique Vicentim. Howie Lim, Ibu Ketty, Jose Garcia, Junming Wu, Ketty Rosenfeld, Luana Araujo, Mariano Ruiz, Maryam Motaghi, Maso, Omar Cherkaoui, Panyaphon Phiphatkunarnon, Pedro Moreno, Poyao Huang, Souad Orhan, Stephane Ku, Tanat Chinbunchorn, Teak Sowaprux, Top Medping, and Yalda Toofan.

## SUPPORTING INFORMATION

**S1 Table.** Access to and actualization of gender-affirming resources among self-identified transgender and non-binary individuals who participated in the COVID-19 Disparities Survey, stratified by country (April 16 – August 3, 2020, N=964)

|  | European<br>Region | South-East Asia<br>Region | Region<br>of the<br>Americas | Eastern<br>Mediterranean<br>Region | Western<br>Pacific<br>Region | African<br>Region | p-value<sup>a</sup> |
| --- | --- | --- | --- | --- | --- | --- | --- |
| **Experienced reduced access to one or more gender affirming resource below<sup>b</sup>** | 116 / 207<br>(56.0%) | 109 / 204<br>(53.4%) | 32 / 53<br>(60.4%) | 30 / 56<br>(53.6%) | 6 / 25<br>(24.0%) | 15 / 23<br>(65.2%) | 0.043 |
| Hormone therapy and/or gender affirming medication | 47 / 116<br>(40.5%) | 55 / 186<br>(29.6%) | 10 / 25<br>(40.0%) | 11 / 32<br>(34.4%) | 4 / 19<br>(21.1%) | 8 / 13<br>(61.5%) | 0.081 |
| Surgical aftercare | 37 / 93<br>(39.8%) | 51 / 175<br>(29.1%) | 7 / 15<br>(46.7%) | 8 / 33<br>(24.2%) | 2 / 18<br>(11.1%) | 8 / 12<br>(66.7%) | 0.008 |
| Cosmetic supplies and services, e.g., makeup, wigs, and hair removal | 68 / 176<br>(38.6%) | 62 / 181<br>(34.3%) | 21 / 46<br>(45.7%) | 21 / 47<br>(44.7%) | 3 / 23<br>(13.0%) | 9 / 15<br>(60.0%) | 0.025 |
| Mental health counseling and therapy* | 62 / 128<br>(48.4%) | 73 / 186<br>(39.2%) | 25 / 42<br>(59.5%) | 12 / 42<br>(28.6%) | 3 / 19<br>(15.8%) | 9 / 18<br>(50.0%) | 0.004 |
| Body modifiers, e.g., binders and packing material | 56 / 134<br>(41.8%) | 58 / 179<br>(32.4%) | 17 / 40<br>(42.5%) | 14 / 46<br>(30.4%) | 3 / 21<br>(14.3%) | 8 / 13<br>(61.5%) | 0.030 |
| **Compared to before the COVID-19 pandemic, how often able to live according to their gender** |  |  |  |  |  |  |  |
| More or a lot more | 25 / 381<br>(6.6%) | 20 / 236<br>(8.5%) | 7 / 84<br>(8.3%) | 6 / 71<br>(8.5%) | 0 / 34<br>(0.0%) | 6 / 33<br>(18.2%) | 0.000 |
| About the same | 181<br>(47.5%) | 156<br>(66.1%) | 53<br>(63.1%) | 26<br>(36.6%) | 24<br>(70.6%) | 13<br>(39.4%) |  |
| Less or not at all | 175<br>(45.9%) | 60<br>(25.4%) | 24<br>(28.6%) | 39<br>(54.9%) | 10<br>(29.4%) | 14<br>(42.4%) |  |

<sup>a</sup> p-values were calculated using chi-squared and Fischer's exact tests as appropriate <sup>b</sup> Denominators excluded participants who were not presented with these questions, did not respond, or said that the resource was not applicable to them

**S2 Table.** Mental health and resiliency indicators among transgender and non-binary individuals who participated in the COVID-19 Disparities Survey, stratified by country (April 16 – August 3, 2020, N=964^a^

|  | European<br>Region | South-East Asia<br>Region | Region<br>of the<br>Americas | Eastern<br>Mediterranean<br>Region | Western<br>Pacific<br>Region | African<br>Region | p-value<sup>b</sup> |
| --- | --- | --- | --- | --- | --- | --- | --- |
| **Screened positive per PHQ-4 (score ≥3)** |  |  |  |  |  |  |  |
| Depression | 232 / 400<br>(58.0%) | 66 / 222<br>(29.7%) | 46 / 82<br>(56.1%) | 48 / 76<br>(63.2%) | 21 / 40<br>(52.5%) | 18 / 32<br>(56.3%) | ≤ 0.001 |
| Anxiety | 392 / 856<br>(45.8%) | 392 / 856<br>(45.8%) | 392 / 856<br>(45.8%) | 392 / 856<br>(45.8%) | 392 / 856<br>(45.8%) | 392 / 856<br>(45.8%) | ≤ 0.001 |
| **Felt lonely since COVID-19 began** |  |  |  |  |  |  |  |
| Yes | 685 / 957<br>(71.6%) | 685 / 957<br>(71.6%) | 685 / 957<br>(71.6%) | 685 / 957<br>(71.6%) | 685 / 957<br>(71.6%) | 685 / 957<br>(71.6%) | 0.063 |
| **Frequency of suicidal ideation since**<br>**COVID-19 vs. 6 months prior** |  |  |  |  |  |  |  |
| Was and remains rare | 648 / 928<br>(69.8%) | 648 / 928<br>(69.8%) | 648 / 928<br>(69.8%) | 648 / 928<br>(69.8%) | 648 / 928<br>(69.8%) | 648 / 928<br>(69.8%) | 0.020 |
| Decreased from frequent to rare | 116<br>(12.5%) | 116<br>(12.5%) | 116<br>(12.5%) | 116<br>(12.5%) | 116<br>(12.5%) | 116<br>(12.5%) |  |
| Increased from rare to frequent | 93<br>(10.0%) | 93<br>(10.0%) | 93<br>(10.0%) | 93<br>(10.0%) | 93<br>(10.0%) | 93<br>(10.0%) |  |
| Was and remains frequent | 71<br>(7.6%) | 71<br>(7.6%) | 71<br>(7.6%) | 71<br>(7.6%) | 71<br>(7.6%) | 71<br>(7.6%) |  |
| **Reported having sources of hope, strength,**<br>**comfort, and peace** |  |  |  |  |  |  |  |
| Yes | 489 / 768<br>(63.7%) | 489 / 768<br>(63.7%) | 489 / 768<br>(63.7%) | 489 / 768<br>(63.7%) | 489 / 768<br>(63.7%) | 489 / 768<br>(63.7%) | ≤ 0.001 |
| **Reported being intent on finding emotional**<br>**support and therapy<sup>c</sup>** |  |  |  |  |  |  |  |
| Agree | 467 / 851<br>(54.9%) | 467 / 851<br>(54.9%) | 467 / 851<br>(54.9%) | 467 / 851<br>(54.9%) | 467 / 851<br>(54.9%) | 467 / 851<br>(54.9%) | ≤ 0.001 |
| Disagree | 136<br>(16.0%) | 136<br>(16.0%) | 136<br>(16.0%) | 136<br>(16.0%) | 136<br>(16.0%) | 136<br>(16.0%) |  |
| **Reported believing they could live a happy,**<br>**full life despite the pandemic<sup>c</sup>** |  |  |  |  |  |  |  |
| Agree | 516 / 847<br>(60.9%) | 516 / 847<br>(60.9%) | 516 / 847<br>(60.9%) | 516 / 847<br>(60.9%) | 516 / 847<br>(60.9%) | 516 / 847<br>(60.9%) | ≤ 0.001 |
| Disagree | 120<br>(14.2%) | 120<br>(14.2%) | 120<br>(14.2%) | 120<br>(14.2%) | 120<br>(14.2%) | 120<br>(14.2%) |  |

<sup>a</sup> Denominators excluded individuals who did not respond or reported not knowing their answer unless otherwise noted <sup>b</sup> p-values were calculated using chi-squared and Fischer's exact tests as appropriate <sup>c</sup> Denominator includes those who stated "neither agree nor disagree"

**S3 Table:** Socioeconomic indicators among transgender and non-binary individuals who participated in the COVID-19 Disparities Survey, stratified by country (April 16 – August 3, 2020, N=964)^a^

|  | European<br>Region | South-East Asia<br>Region | Region of<br>the<br>Americas | Eastern<br>Mediterranean<br>Region | Western<br>Pacific<br>Region | African<br>Region | p-value<sup>b</sup> |
| --- | --- | --- | --- | --- | --- | --- | --- |
| **Lost job due to COVID-19** | 74 / 446<br>(16.6%) | 38 / 238<br>(16.0%) | 13 / 86<br>(15.1%) | 15 / 83<br>(18.1%) | 5 / 40<br>(12.5%) | 2 / 35<br>(5.7%) | 0.614 |
| **Expected reduction in income** |  |  |  |  |  |  |  |
| 0% | 113 / 436<br>(25.9%) | 39 / 340<br>(16.3%) | 25 / 86<br>(29.1%) | 19 / 79<br>(24.1%) | 9 / 40<br>(22.5%) | 9 / 35<br>(25.7%) | 0.055 |
| 1-39% | 123<br>(28.2%) | 66<br>(27.5%) | 20<br>(23.3%) | 20<br>(25.3%) | 14<br>(35.0%) | 11<br>(31.4%) |  |
| 40-99% | 140<br>(32.1%) | 110<br>(45.8%) | 34<br>(39.5%) | 25<br>(31.7%) | 13<br>(32.5%) | 10<br>(28.6%) |  |
| 100% | 60<br>(13.8%) | 25<br>(10.4%) | 7<br>(8.1%) | 15<br>(19.0%) | 4<br>(10.0%) | 5<br>(14.3%) |  |
| **Expected to lose health insurance<sup>c</sup>** |  |  |  |  |  |  |  |
| Yes | 32 / 307<br>(10.4%) | 42 / 165<br>(25.5%) | 11 / 67<br>(16.4%) | 16 / 51<br>(31.4%) | 7 / 35<br>(20.0%) | 1 / 21<br>(4.8%) | < 0.001 |
| No | 203<br>(66.1%) | 94<br>(57.0%) | 48<br>(71.6%) | 27<br>(52.9%) | 18<br>(51.4%) | 12<br>(57.1%) |  |
| **Receipt and need of financial aid** |  |  |  |  |  |  |  |
| Received and not needed | 10 / 375<br>(2.7%) | 6 / 208<br>(2.9%) | 2 / 75<br>(2.7%) | 3 / 64<br>(4.7%) | 0 / 32<br>(0.0%) | 0 / 30<br>(0.0%) | <0.001 |
| Not received but not needed | 86<br>(22.9%) | 29<br>(13.9%) | 19<br>(25.3%) | 13<br>(20.3%) | 6<br>(18.8%) | 5<br>(16.7%) |  |
| Received and needed | 53<br>(14.1%) | 92<br>(44.2%) | 23<br>(30.7%) | 8<br>(12.5%) | 8<br>(25.0%) | 5<br>(16.7%) |  |
| Not received and needed | 226<br>(60.3%) | 81<br>(38.9%) | 31<br>(41.3%) | 40<br>(62.5%) | 18<br>(56.3%) | 20<br>(66.7%) |  |
| **Had cut or reduced meals** | 167 / 420<br>(39.8%) | 82 / 230<br>(35.7%) | 28 / 82<br>(34.2%) | 45 / 75<br>(60.0%) | 12 / 37<br>(32.4%) | 16 / 34<br>(47.1%) | 0.004 |
| **Among those employed,**<br>**ability to miss work** |  |  |  |  |  |  |  |
| Was telecommuting or on paid leave | 81 / 214<br>(37.9%) | 40 / 127<br>(31.5%) | 17 / 42<br>(40.5%) | 11 / 34<br>(32.4%) | 6 / 21<br>(28.6%) | 6 / 20<br>(30.0%) | 0.165 |
| Cannot afford to miss work but was<br>following confinement orders | 72<br>(33.6%) | 34<br>(26.8%) | 9<br>(21.4%) | 9<br>(26.5%) | 5<br>(23.8%) | 10<br>(50.0%) |  |
| Cannot afford to stay home and must<br>work to survive | 61<br>(28.5%) | 53<br>(41.7%) | 16<br>(38.1%) | 14<br>(41.2%) | 10<br>(47.6%) | 4<br>(20.0%) |  |

<sup>a</sup> Denominators excluded individuals who did not respond or reported not knowing their answer unless otherwise noted <sup>b</sup> p-values were calculated using chi-squared and Fischer's exact tests as appropriate <sup>c</sup> Denominator includes those who reported they "might or might not" lose their health insurance

